# Estimating tuberculosis-related patient costs in KwaZulu-Natal, South Africa

**DOI:** 10.64898/2026.05.18.26353472

**Authors:** Ian Yoon, Indira Govender, Palwasha Khan, Mareca Sithole, Nicky McCreesh, Alison D Grant, Sedona Sweeney

**Author notes:** Ethical Approval: The study was approved in South Africa by the Biomedical Research Ethics Committee, University of KwaZulu-Natal, Durban, South Africa under the reference number BF472/19. Ethical approval was also obtained from The London School of Hygiene & Tropical Medicine under the reference number 17728.

## Abstract

**Summary:** In a cross-sectional study, we calculated direct and indirect costs incurred by people prior to starting tuberculosis (TB) treatment in primary healthcare facilities in KwaZulu-Natal, South Africa. We related the total costs to patient income to explore the economic impact of TB care-seeking and contribute to the literature by exploring differences between those with and without TB symptoms.

**Background:** Patient costs during tuberculosis (TB) treatment in South Africa are high. There are fewer data about the costs incurred prior to starting treatment. We measured pre-TB treatment costs for people in rural KwaZulu-Natal, South Africa.

**Design/methods:** In the context of a TB case-contact study, we interviewed people starting TB treatment at primary healthcare facilities in rural South Africa. We estimated total direct and indirect costs incurred by respondents and their households in the three months prior to starting TB treatment. We estimated other coping costs, such as selling productive assets, as well as the value of any loans taken.

**Results:** Among 98 participants (52 female, median age 36 years), 86/98 (88%) reported one or more symptoms from the WHO 4-symptom TB screening tool prior to starting treatment. The median total pre-treatment cost for TB affected households was USD 10.78 (IQR: [4.13 – 20.23]). Total, pre-treatment costs for those with TB symptoms were USD 10.78 (IQR: [4.83 – 20.23]) compared to USD 8.91 (IQR: [1.27 – 22.19]) for those without TB symptoms.

**Conclusions:** Whilst TB testing and care is free in South African public health facilities, patients still face costs that are burdensome. Our results indicate people affected by TB, including patients and their families, also face an economic burden. Our study highlights the need for further consideration of social protection policies to reduce the economic effects of asymptomatic TB.

**Key Messages:** While TB testing and treatment are free in South African public health facilities, patients still face significant financial costs related to TB care-seeking. There is limited evidence on the economic costs incurred prior to starting treatment, particularly regarding the difference between people with and without TB symptoms.

People without TB symptoms faced similar pre-treatment costs compared to those with TB symptoms.

In rural South Africa, reducing pre-treatment costs for those without TB symptoms may justify active case-finding initiatives to identify these people earlier.

## Introduction

In 2024, the WHO estimates that 10.7 million people developed tuberculosis disease (TB) globally and 1.23 million people died from TB-related illness. Diagnosis and treatment of TB have improved due to advances, for example, in case-finding strategies, diagnostic platforms, and treatment regimens. However, there remains a gap of three million people between estimated incident TB and those who start TB treatment. The WHO highlights that closing this gap will require increased case-detection including those who do not have TB symptoms or seek care (World Health Organization, 2025).

The South African National Strategic Plan on HIV, TB and Sexually Transmitted Infections 2023-2028 highlights the importance of TB screening among adult clinic attendees. This includes screening all adults attending clinic for TB symptoms using the WHO 4-symptom screening tool (cough, weight loss, night sweats or fever). Additionally, targeted universal testing for TB (TUTT) aims to identify people at higher risk of active TB through annual sputum testing of people with increased risk, regardless of reported symptoms (South African National Department of Health, 2023) (including people living with HIV (PLHIV), people treated for TB in the previous two years, and close contacts of people with TB).

Improved case finding for TB is often cited as an important step in achieving the goal of “eliminating catastrophic costs for TB-affected households” articulated through the World Health Organization’s End TB Strategy (World Health Organization, 2015). A 2023 systematic review of TB-related costs showed that across 135 low- and middle-income countries (LMIC), over half of people with TB incurred costs at a catastrophic level (greater than 20% of household income), and roughly half of these costs occurred before treatment start. Costs related to TB have been shown to create access barriers to treatment, ultimately worsening health outcomes and increasing disease transmission (Portnoy et al., 2023). In South Africa, although TB testing and treatment are free under national policy guidelines (South African National Department of Health, 2023), most people with TB (56.2%) still encounter catastrophic costs due to TB, largely driven by income loss (68% of total patient costs) (South African National Department of Health, 2024).

From a patient cost perspective, there may be benefits to people for earlier TB case detection, both before and after treatment initiation. This may be particularly true for people with intermittent TB symptoms, who may otherwise be missed in routine TB case-detection. There is limited evidence concerning pre-treatment costs incurred by with asymptomatic TB. Our aim was to describe pretreatment costs for people treated for TB in South Africa and compare costs incurred by people who reported TB symptoms at TB treatment start against those incurred by people who reported no symptoms.

## Methods

### Study Design and Setting

The setting for this study is public primary healthcare clinics serving the Health and Demographic Surveillance site (HDSS) of the African Health Research Institute (AHRI). This is a long-established surveillance area which covers 845⍰km^2^ and includes a population of approximately 140,000 individuals across 20,000 households (Gareta et al., 2021).

#### Study Population

Ours is a sub-study of a wider TB household case-contact study, the Africa Health Research Institute (AHRI) Household Contact Study (AHCoS) study. This study recruited index people aged 15 years or older with bacteriologically-confirmed TB starting treatment at a health facility serving the HDSS and living in a household with at least one child aged 2-14 years. The study design is described in full elsewhere (Khan et al., 2024).

Between October 2022 and January 2023, consecutive AHCoS enrollees were approached by telephone to participate in this costing study. All study participants were asked about their TB-related costs for due to TB in three months prior to starting TB treatment.

### Data Collection

Our questionnaire was developed using the guidance issued in “Tuberculosis patient cost surveys: a handbook” developed by the World Health Organization (World Health Organization, 2017). We collected demographic characteristics such as age, gender, and employment status. The questionnaire included questions regarding the visits made by the individual within the three months prior to starting TB treatment. We collected details regarding the number of visits made to a range of providers (including primary health centres, mobile primary health centres, pharmacies, private doctors, and traditional health practitioners), the reason for the visit, time spent travelling to and at the facility during those visits, and any direct costs incurred from travel to or at the facility. In addition, we asked about coping mechanisms that people used to manage the cost of illness. This included selling assets, using savings, receiving gifts, and taking loans. Each person participating in the study was interviewed once over the period March 2023 to February 2024.

We estimated direct and indirect costs incurred by study participants and their households. Direct costs included payment for accommodation, consultant fees, medicines, food, diagnostic fees for scans, tests and x-rays, procedures, travel and all other medical costs. Our primary measure of indirect costs is estimated using a human capital approach. We include the opportunity cost of time spent on care-seeking both by the person with TB and any guardians or informal carers for that person. Time was valued using individual income, which was directly elicited through the questionnaire. To support comparability with other studies, we also report on time (minutes) spent on care-seeking or away from work for both caregivers and survey participants.

### Data analysis

We included all visit costs where the reported reason for visit was potentially related to TB or TB symptoms. This included feeling sick and referrals from other service providers. We excluded the cost of visits for other unrelated health issues, including HIV and antenatal care.

For people who were employed on a full-time basis, income was converted into a per-minute rate accounting for public holidays, annual leave, and sick leave. For those who were employed part-time, their per-minute rate estimates were based on their working patterns prior to starting TB treatment. We did not directly collect the incomes of guardians and carers. Therefore, we estimate their income loss as the per-minute rate equivalent to the individual with TB who they accompanied multiplied by time (measured in minutes) spent travelling to or at the clinic.

Data collection was completed by a single research assistant to maintain consistency in asking questions. The research assistant was an employee of AHRI. The questionnaire was administered via telephone. Data were collected and entered at the time of interview by the data enumerator using surveys captured in a secure electronic database (Harris et al., 2009). Data cleaning, processing, and statistical analysis were completed using Stata 14 for our “available case” analysis in which data are assumed missing completely at random (StataCorp, 2025).

We defined people with symptomatic TB as those starting treatment who had bacteriologically-confirmed TB based on a positive result from a sputum specimen (either Xpert MTB/RIF Ultra or culture-positive for *M. tuberculosis*) who reported one or more symptoms from the WHO screening tool at the point of starting treatment. We defined people with asymptomatic TB as those who did not report TB symptoms at AHCoS enrollment (World Health Organization, 2024). We report descriptive statistics for the characteristics of study participants as well as their health-seeking behaviour and related costs. To manage non-normality of cost data as well as skewness, we report both the mean and median of all costs, along with the standard deviation and inter-quartile range. All reported costs were converted from the South African Rand to the United States Dollar at the average rate of conversion during the period of data collection March 2023 to February 2024: 18.67 ZAR = 1 USD (www.Oanda.com, 2024).

We used a concentration index to estimate socioeconomic inequality in pre-treatment costs and reported incomes. We calculated concentration index values and the corresponding p-values to determine statistical significance using Stata (StataCorp, 2025),

## Ethical Considerations

The study was approved in South Africa by the Biomedical Research Ethics Committee, University of KwaZulu-Natal, Durban, South Africa under the reference number BF472/19. Ethical approval was also obtained from The London School of Hygiene & Tropical Medicine under the reference number 17728. All individuals at least 18 years of age gave written or witnessed verbal consent to participate in the study.

## Results

Table 1 - Characteristics of study populationWe attempted to contact 124 individuals. Of these, 24 could not be reached or declined to take part, and one was enrolled incorrectly (subsequently determined not to have bacteriologically-confirmed TB), leaving 98 people in our analysis.

**Table 1.** Characteristics of study population by TB symptom status.

|  | All<br>n=98 | TB-asymptomatic<br>n=12 | TB-symptomatic<br>n=86 |
| --- | --- | --- | --- |
| **Gender** |  |  |  |
| Female | 52 (55%) | 5 (41%) | 45 (53%) |
| Male | 46 (47%) | 7 (58%) | 41 (47%) |
| **Median age in years [IQR]** | 36 [29 – 50] | 38 [26 – 51] | 36 [29 - 50] |
| **Highest level of educational attained (%)** |  |  |  |
| None | 7 (7%) | 0 (0%) | 7 (8%) |
| Primary | 19 (19%) | 4 (33%) | 15 (17%) |
| Secondary | 69 (71%) | 8 (67%) | 61 (71%) |
| Post-Matric | 3 (3%) | 0 (0%) | 3 (3%) |
| **Employment Status at time of survey (%)** |  |  |  |
| Not Employed | 70 (72%) | 10 (84%) | 60 (70%) |
| Part-Time Employed | 12 (12%) | 1 (8%) | 11 (13%) |
| Full Time Employed | 15 (15%) | 1 (8%) | 14 (16%) |
| N/A<sup>1</sup> | 1 (1%) | 0 (0%) | 1 (1%) |
| **Median Annual Income USD 2024 [IQR]** | 1,478 [309 – 2,571] | 1,607 [643 – 1,928] | 1,279 [309 – 2,250] |

<sup>1</sup> N/A: Other (not specified)

Among 98 participants, the median age was 36 years (IQR: [29-50]), 52 (55%) were female, 65 (66%) were HIV-positive and 86 (88%) reported one or more TB symptoms at the time of treatment initiation (Table 1). Most participants had some secondary education, and the annual mean income level was USD 80.34 per year. Across all health provider types, most people went to a primary health centre.

Table 2 shows the descriptive pre-treatment costs for all participants disaggregated by TB-symptom status. For all study participants, the median total pre-treatment cost to the individual was USD 10.78 (IQR: [4.13 - 20.23]). The median direct cost was USD 4.28 (IQR: [2.14 – 14.46]). The median indirect cost was USD 2.37 (IQR: [0.29 - 5.76]).

**Table 2.** Pre-treatment TB-related costs by TB symptom status (USD 2024) ^2^.

|  | All<br>n=98 |  | TB-asymptomatic<br>n=12 |  | TB-symptomatic<br>n=86 |  | U-Test<br>p-value | Concentration<br>Index<br>(p-value) |
| --- | --- | --- | --- | --- | --- | --- | --- | --- |
|  | Mean (SD) | Median [IQR] | Mean (SD) | Median [IQR] | Mean (SD) | Median [IQR] |  |  |
| **Total Costs** | **20.06 (38.1)** | **10.78 [4.13 - 20.23]** | **15.12 (18.17)** | **8.91 [1.27 - 22.19]** | **20.74 (40.1)** | **10.78 [4.83 - 20.23]** | **0.45** | **0.14 (0.21)** |
| **All Direct Costs** | **15.76 (33.76)** | **4.28 [2.14 - 14.46]** | **12.45 (15.32)** | **6.96 [1.07 - 17.27]** | **16.22 (35.59)** | **4.28 [2.14 - 14.46]** | **0.82** | **0.02 (0.89)** |
| Travel Costs | 5.09 (10.78) | 2.36 [1.29 - 5.36] | 3.19 (3.49) | 2.41 [0 - 5.36] | 5.35 (11.41) | 2.36 [1.61 - 5.36] | - | 0.09 (0.46) |
| Consultation Costs | 2.75 (8.75) | 0 [0 - 0] | 2.59 (6.2) | 0 [0 - 0] | 2.78 (9.07) | 0 [0 - 0] | - | -0.19 (0.31) |
| Laboratory Costs | 0 (0) | 0 [0 - 0] | 0 (0) | 0 [0 - 0] | 0 (0) | 0 [0 - 0] | - | - |
| Other Services | 0.24 (2.03) | 0 [0 - 0] | 0 (0) | 0 [0 - 0] | 0.28 (2.17) | 0 [0 - 0] | - | -0.62 (0.02) |
| Drugs | 4.69 (16.59) | 0 [0 - 0] | 4.33 (6.73) | 0 [0 - 9.64] | 4.74 (17.54) | 0 [0 - 0] | - | 0.18 (0.38) |
| Food | 2.16 (3.53) | 1.07 [0 - 2.68] | 1.72 (1.67) | 1.07 [0.54 - 2.81] | 2.22 (3.71) | 1.07 [0 - 2.68] | - | -0.14 (0.15) |
| Accommodation | 0.83 (1.69) | 0 [0 - 0] | 0.62 (1.48) | 0 [0 - 0] | 0.85 (1.72) | 0 [0 - 1.07] | - | -0.09 (0.45) |
| **Indirect Costs** | **4.3 (7.12)** | **2.37 [0.29 - 5.76]** | **2.68 (3.56)** | **0.25 [0.07 - 5.94]** | **4.52 (7.47)** | **2.4 [0.81 - 5.61]** | **0.21** | **.52 (0.00)** |
| Seeking Care - Person with TB | 4.24 (7.05) | 2.21 [0.24 - 5.67] | 2.61 (3.54) | 0.24 [0 - 5.88] | 4.46 (7.39) | 2.39 [0.81 - 5.61] | - | 0.56 (0.00) |
| Seeking Care - Carer | 0.06 (0.14) | 0 [0 - 0.07] | 0.07 (0.1) | 0.01 [0 - 0.14] | 0.06 (0.15) | 0 [0 - 0.06] | - | 0.24 (0.10) |
| Travel Time - Person with TB | 234.55 (225.41) | 180 [120 - 255] | 186.25 (169.84) | 180 [20 - 300] | 241.21 (232.04) | 180 [120 - 240] | - | -0.07 (0.22) |
| Travel Time - Carer | 66.87 (152.17) | 0 [0 - 110] | 70.83 (106.64) | 10 [0 - 135] | 66.32 (157.89) | 0 [0 - 110] | - | -0.21 (0.87) |
| **Total Visits** | **1.66 (1.10)** | **1.00 [1.00 - 2.00]** | **1.45 (0.78)** | **1.00 [1.00 - 1.50]** | **1.69 (1.10)** | **1.00 [1.00 - 2.00]** | **-** | **-** |

<sup>2</sup> USD: United States Dollar; SD: Standard Deviation; IQR: Inter Quartile Range

**Table 4.** Coping mechanisms and effects of TB by TB symptom status.

|  |  | All<br>n=98 | TB-symptomatic<br>n=86 | TB-asymptomatic<br>n=12 |
| --- | --- | --- | --- | --- |
| **Coping mechanism**<br>**Frequency (USD**<br>**2024 Value [IQR])** | **Drew on savings** | 3 (80 [80 - 107]) | 2 (94 [80 – 107]) | 1 (80) |
|  | **Took a loan** | 21 (54 [27 - 80]) | 20 (48 [27 – 80]) | 1 (80) |
|  | **Sold an asset** | None | None | None |
|  | **Received a gift** | 12 (43 [19 - 67]) | 10 (43 [21 - 80]) | 2 (35 [16 - 54]) |
| **Status after TB (%**<br>**of Participants)** | **Worse-off** | 26 (27%) | 25 (30%) | 1 (8%) |
|  | **Same** | 72 (73%) | 61 (70%) | 11 (92%) |
|  | **Better** | None | None | None |
| **Reduced work due**<br>**to TB (% of**<br>**Participants)** | **Yes** | 13 (13%) | 11 (13%) | 2 (17%) |
|  | **No** | 27 (27%) | 25 (29%) | 2 (17%) |
|  | **N/A<sup>3</sup>** | 58 (60%) | 50 (58%) | 8 (66%) |
| **Lost income due to**<br>**TB (% of**<br>**Participants)** | **Yes** | 3 (3%) | 3 (4%) | None |
|  | **No** | 10 (10%) | 8 (9%) | 2 (17%) |
|  | **N/A** | 85 (87%) | 75 (87%) | 10 (83%) |

<sup>3</sup> N/A - Not employed prior to TB treatment initiation

**Table 5.** Shapiro-Wilk Normality Robustness Tests.

| Variable | Number of Observations | W (Test Statistic) | Z (Standardized Statistic) | Significance (p-value) |
| --- | --- | --- | --- | --- |
| Indirect Costs | 98 | 0.54279 | 8.009 | 0.00 |
| Direct Costs | 98 | 0.5794 | 7.824 | 0.00 |
| Total Cost | 98 | 0.59572 | 7.736 | 0.00 |

We find that individuals with TB symptoms had greater median total costs than costs for those without TB symptoms. However, the IQRs for each are overlapping between groups. Median total TB-related costs for people with symptomatic TB were USD 10.78 (IQR: [4.83 - 20.23]) compared to median total costs of USD 8.91 (IQR: [1.27 – 22.19]) for those with asymptomatic TB. We could not find any evidence for a difference between those with and without TB symptoms in median total, direct, and indirect costs (Table 2).

For direct costs, we find these were lower for people with symptomatic TB (USD 4.28 (IQR: [2.14 – 14.46]) compared to those with asymptomatic TB (USD 6.96; IQR: [1.07 – 17.27]). Regardless of TB symptom status, direct costs are composed of travel and food costs.

Median indirect costs for people with symptomatic TB were USD 2.40 (IQR: [0.81 - 5.61]). For people with asymptomatic TB, median indirect costs were USD 0.25 (IQR: [0.07 - 5.94]). For both groups, indirect costs are driven by time lost for the individual with TB seeking TB-related care.

Finally, we find that for both people with and without TB symptoms, median total number of health facility visits was 1.00, though the IQR for those with symptomatic TB was wider than for those with asymptomatic TB.

We assess the distribution of pre-treatment financial burden across socioeconomic status via a concentration index. We find no statistically significant inequality in the overall distribution of total costs (Concentration Index = 0.14, p = 0.21) or total direct costs (Concentration Index = 0.02, p = 0.89). However, indirect costs are concentrated towards wealthier people (Concentration Index = 0.52, p < 0.01).

### Economic wellbeing impact and coping strategies

#### Error! Reference source not found

3 presents pre-treatment costs as a proportion of annual income for respondents who reported income. Pre-treatment costs were a small proportion of annual income for both populations. However, costs as a proportion of income decrease related to wealth quartile.

35% of all TB-affected households employed a coping strategy to manage the cost of care-seeking. The most common strategy was taking a loan (21%) with a median value of USD 53.56 (IQR: [26.78 – 80.34]). People also reported receiving gifts from family and friends (12%) with a median value of USD 42.85 (IQR: [18.75 – 66.95]). Most people interviewed stated that they were no worse off economically due to TB (73%). For those who were employed prior to being diagnosed with TB, 30% had to reduce work. However, only 23% of those individuals stated that they had lost income. There was limited variation in wellbeing and coping strategies across subgroups.

## Discussion

We find that pre-treatment costs for people with TB range from USD 4.13 to 20.23. Total costs are largely driven by direct costs, particularly travel and food. People with asymptomatic TB face similar pre-treatment TB-related costs to those encountered by people with symptomatic TB. Our data suggest this may be because people with asymptomatic TB still pay directly for travel to a health facility and indirectly through lost income.

Our definition of asymptomatic TB is dependent on the results of a TB symptom screen at the time of treatment initiation, rather than ‘during screening’ as per the WHO definition (World Health Organization, 2024). Previous research suggests that for many people with TB, symptoms may be mild or intermittent – perhaps significant enough to warrant visiting a health facility, but not identifiable as TB through a clinical screening. Kendall and colleagues add that existing data already show that these individuals likely account for a meaningful fraction of *M. tuberculosis* transmission and that large-scale, active case-finding is required to reach global targets for reduction in TB incidence (Kendall et al., 2021; McCreesh et al., 2026). Our findings extend that a better understanding of the range of potential presentation of TB symptoms is important for improving social protection in addition to clinical reasons.

Compared to Foster et al., who explicitly reported pre-treatment costs, we find larger median direct and indirect costs for this population (Foster et al., 2015). In addition, we find that direct costs compose a greater proportion of pre-treatment costs. (Foster et al., 2015; Mudzengi et al., 2017; Tanimura et al., 2014). We identify two reasons for this difference. Firstly, low wages in uMkhanyakude district relative to the national average imply limited productivity losses in a human capital approach (South African National Department of Cooperative Governance and Traditional Affairs, 2020; South African National Department of National Treasury, 2021). Secondly, high unemployment in the study group compared to district average would also limit productivity losses in the human capital approach. In addition, we find that direct costs are lower as a proportion of income compared to the literature which may also be attributable to low wages and/or high unemployment (Foster et al., 2015).

Our estimates of pre-treatment TB-related costs are consistent with other South African patient cost studies that find costs related to TB care-seeking to be non-catastrophic in themselves (Foster et al., 2015; Ramma et al., 2015; Tanimura et al., 2014). This does not preclude catastrophic costs from the entire TB episode. As Foster and colleagues show in South Africa, more than half of total TB-related patient costs are incurred after starting TB treatment and well above 10% of median annual income in our sample (Foster et al., 2015).

However, even non-catastrophic costs can cause TB-affected households to employ coping strategies that reduce wealth like selling assets, borrowing money from family, and taking loans. Employment of coping strategies early in the TB episode can potentially be damaging for household economic wellbeing long-term, even if costs do not exceed the threshold for catastrophic costs. This can include lower economic productivity from selling assets. Moreover, if coping strategies affect household liquidity, such as from loan interest payments or paying borrowed money, this can result in a reduction in nutrition or educational attainment if families can no longer afford school fees (Saqib et al., 2018).

Finally, our analysis shows a significant concentration of indirect pre-treatment costs among wealthier people with TB. This follows from our human capital approach used to value lost time; wealthier individuals are more likely to have formal employment and higher hourly wages, and the monetary value of their time spent seeking care is higher. However, while the poorest people with TB may incur lower absolute indirect costs, the opportunity cost of their lost time may have a larger impact on their livelihoods than for wealthier counterparts.

## Limitations

To increase the sample size, data enumerators made significant efforts to reach the study population. However, our study was of a small sample of those with TB in a specific population recruited through index identification. This means that the sample may not be representative of the wider population attending health clinics.

We asked individuals to recall health expenditures in the last three months. While this is within the recommended period, it is possible that people may inaccurately recall the time spent and expenses incurred related to TB care seeking. We highlight that our analysis is a localised estimate and would caution against generalisation (Gunda et al., 2022).

Missing income data also presents a challenge for estimating the magnitude of TB-related costs as well as determining the extent of catastrophic health expenditure. Given our small sample size, we could not omit individuals who did not report income. If poorer individuals were disproportionately less likely to report income, then our estimates of indirect costs will be biased upwards (Kerr, 2025). Therefore, we followed several studies in completing a separate analysis using an imputed income of USD 1.00 (Ataguba and McIntyre, 2012; Foster et al., 2015).

People in the wider study represented ambulant clinic attendees and may therefore exclude people treated as inpatients, as well as people not attending clinics at all. While this may have affected our pre-treatment cost estimates, these individuals also may be of lower socio-economic status as compared to people from urban areas, and therefore any pre-treatment costs would represent a greater proportion of their household income (Clarke-Deelder et al., 2022). In addition, people attending ambulant care may be experiencing less severe symptoms of TB as compared to those treated as inpatients.

Methodologically, there is no consensus in the literature regarding the measurement of indirect costs. We chose our primary measure as income lost to support comparability with other studies, though would echo calls that further methodological research on the measurement of indirect costs is needed (Mudzengi et al., 2017; Sweeney et al., 2020).

## Policy Implications and Future Research

We found that both symptomatic and asymptomatic people with TB face pre-treatment costs, and there was no significant difference in costs incurred by the two groups. Our results support active case-finding initiatives to identify people with asymptomatic TB. Increased attention to identifying individuals with TB can not only reduce the incidence of TB in South Africa but also improve social protection for those with TB disease. Specifically, our results emphasize the need for increased coverage of social protection funds to ensure that all TB-affected households are supported. While individuals unable to work due to TB already qualify for public disability grants in South Africa if they prove that they are “unfit to work” for a minimum of six months, this could preclude individuals with mild or transient symptoms. This is important in uMkhanyakude district as it is considered one of the most economically deprived districts in KwaZulu-Natal province and one of the poorest in South Africa (Patrick, 2021). Thus, any cost faced by people with TB may have a greater impact on financial stability or result in reduced care seeking to avoid financial loss. However, grant coverage for people with TB is low (5% of people with drug sensitive TB), so it is unclear to what extent an expanded inclusion criteria would provide coverage to people with asymptomatic TB (Foster et al., 2015).

In the context of reductions in financing for global public health more broadly, our analysis highlights the need for three areas of further research regarding the costs incurred by individuals of different sub-groups. The first is whether identifying people with asymptomatic TB or through their routine ART visit is important for sizing social protection budgets for TB-affected households. The second is if reduced costs incurred by people with TB justify bacteriological testing of individuals who are TB asymptomatic or attending their routine ART visit. The third is if testing strategies required for identification of people with asymptomatic TB or during their routine ART visit is cost-effective from a societal perspective.

## Conclusion

Our study estimates the costs incurred by individuals affected by TB and their households prior to starting TB treatment in uMkhanyakude district, KwaZulu-Natal, South Africa. Our data support the current focus on identification and early treatment of people with asymptomatic TB as we find little difference in pre-treatment costs for people with or without TB symptoms at the time of TB treatment initiation. We show that despite free provision of TB-related care in South Africa, some TB-affected households still face catastrophic care-seeking costs. We show this is especially true amongst poor households, suggesting further social protection programmes targeting these households would have equity benefits.

## Data Availability

All data produced in the present study are available upon reasonable request to the authors

## Supplement

